# Long-Term follow up of Myocardial Function in VA-ECMO

**DOI:** 10.1101/2023.04.11.23288438

**Authors:** Cheng-Ta Yang, Yu-Ting Cheng, Yi-Hsin Chan, Victor Chien-Chia Wu, Dong-Yi Chen, Kuo-Chun Hung, Fu-Chih Hsiao, Ying-Chang Tung, Chia-Pin Lin, Pao-Hsien Chu, Shao-Wei Chen

## Abstract

**Objective:** There is limited evidence regarding the association between myocardial function and long-term survival rate in patients who reach hospital discharge. This study aimed to investigate the association between myocardial function parameters collected at different times from weaning to long-term follow-up and the long-term mortality rate.

**Method:** A cohort of 403 patients successfully weaned from VA-ECMO was identified from a total of 1300 patients who underwent VA-ECMO between 2000-2018 after applying exclusion criteria for age and indications not of interest in the Chang Gung Memorial Hospital Research Database. A retrospective analysis was performed to investigate the effect of ejection fraction timing on long-term mortality.

**Results:** Percentile improvement in EF between ECMO placement and successful weaning is significantly associated with lower cumulative mortality, while the EF value before discharge was significantly associated with better survival. Lastly, the association of lower long-term mortality with EF change from discharge to midterm follow-up and the maximum EF at midterm follow-up was found to be non-significant.

**Conclusions:** This is the first study to provide a comprehensive analysis of echo-cardiographic parameters collected at different times and long-term cumulative mortality in patients who survived VA-ECMO. Improvements in cardiac function and better baseline cardiac function are associated with lower long-term mortality.

## Introduction

Extracorporeal membrane oxygenation (ECMO) is used to provide hemodynamic stability in patients with severe cardiopulmonary dysfunction. Veno-arterial ECMO has been increasingly implemented during extracorporeal cardiopulmonary resuscitation in emergency settings as first-line circulatory support **(Chen, Yih-Sharng, et al, 2008),** and as a bridge to myocardial recovery **(Sertic et al. 2020)** in cardiogenic shocks and postcardiotomy shocks.

By providing oxygenated blood flow to the failing heart, VA-ECMO allows hemodynamic stability and influences left ventricular function over the course of intervention. Basal LV function during weaning plays an important role in determining myocardial recovery and mid-term survival. (**Sertic, Chavez et al. 2021**) **(Pappalardo et al, 2015)** However, the role of the dynamic changes of ejection fraction during VA-ECMO support remain to be investigated. Moreover, there are limited data on the long-term follow-up of patients who survive to discharge due to high mortality and loss of follow-up. While **Wu et al**. previously identified a significantly worse short-to-mid-term survival in postcardiotomy shock resuscitation with post-operative LVEF less than 30% using the database from our medical center, there are no further data on patients with other indications. In addition, despite identifying an association between EF>30 at weaning and significantly improved survived-to-discharge outcomes **(Sertic, Chavez et al. 2021),** there is no current data on a more detailed dynamic change in ejection fraction at weaning, before discharge, and during follow-up for patients with indications from postcardiotomy shock, extracorporeal cardiopulmonary resuscitation (ECPR), and cardiogenic shock. Furthermore, there is a lack of evidence regarding the association between these parameters and long-term mortality.

We aimed to provide a comprehensive analysis of the effects of changes in myocardial function and its absolute baseline values at different time points on the long-term outcome, with the goal of improving the clinical use of ejection fraction for physicians guiding both weaning strategies and managing follow-up in patients who survive to discharge. This paper mainly focuses on VA-ECMO, myocardial function at different times, and long-term survival outcomes.

## Materials and Methods

### Data Source

This retrospective cohort study utilized data collected from the Chang Gung Research Database (CGRD) of the Chang Gung Memorial Hospital (CGMH). Founded in 1976, it has since expanded to oversee three medical centers and four regional hospitals with over 9000 beds and 30000 outpatients daily, one of the largest medical systems in Asia. The multi-institutional electronic medical record database has been used for real-world retrospective studies in Taiwan, containing more pathological and laboratory results than the national insurance database. (**Hsing et al. 2015).** Encrypted digital medical records have been in use for over 20 years and are unlabeled for research use while protecting patients privacy. Informed consent was obtained from the institutional review board of CGMH (approval number: 202100124B).

### Patient identification

After examining the operation notes (surgical reports) between 2003 and 2018 in the CGRD, 1,300 patients administered VA-ECMO were identified. We excluded patients under 18 years of age (n=127), those who received heart transplant or ventricular assist device (n=22) during admission, and those with indications for diseases other than primary cardiac disease (n=252), including septic shock, pulmonary embolism, social indications, and others. (**Figure 1**). Indications were judged based on operation notes and discharge notes. Therefore, 899 patients with indications of interest were included, including post cardiotomy shock, myocarditis, acute myocardial infarction (AMI), ventricular tachycardia (VT) or ventricular fibrillation (VF), decompensated heart failure, and extracorporeal cardiopulmonary resuscitation (ECPR). The remaining 899 patients were classified into the weaned (n=624) and failed weaning (n=275) groups. The time frame for successful weaning was set to a survival of more than 24 hours after weaning to decannulation. Among the 624 weaned patients, 403 survivors were included in further analysis of LVEF (**Figure 1**).

**Figure 1.**
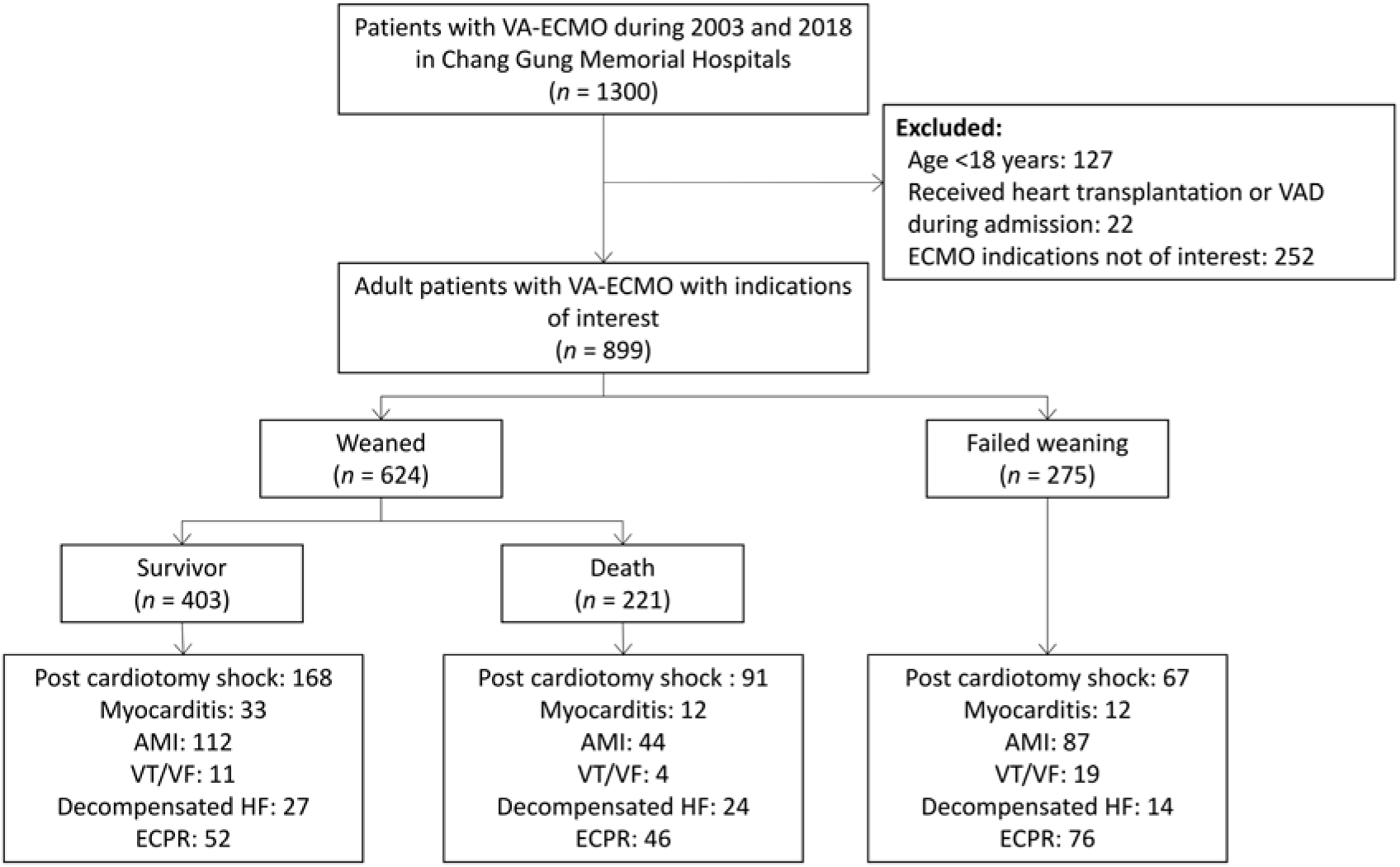
The flowchart for the inclusion and exclusion of the study patients. VA, venoarterial; ECMO, extracorporeal membrane oxygenation; AMI, acute myocardial infarction; VT, ventricular tachycardia; VF, ventricular fibrillation; HF, heart failure; ECPR, extracorporeal cardiopulmonary resuscitation.

### Ejection fraction and outcome

The study outcome was all-cause mortality after discharge. Patients were followed from the day of discharge to the day of death or December 31, 2018, whichever came first. The exposure variables in this study were LVEF according to different definitions. The first definition was improvement from the perioperative period (before ECMO insertion) to before discharge (n=102). The second definition was LVEF before discharge (N=238). The third definition was improvement from discharge to the 180^th^ day after discharge (N=88). The fourth definition was LVEF as the 180^th^ day after discharge. Echocardiography closest (± 90 days) to the 180^th^ day after discharge was selected for analysis.

### Covariates

Covariates included baseline characteristics and ECMO-related features. Baseline characteristics include demographics (age, sex, body mass index, and smoking), comorbidities (diabetes, hypertension, and 14 others), Charlson comorbidity index (CCI) total score, and preoperative laboratory data (hemoglobin, serum creatinine, and 9 others). ECMO-related features include indications, duration from admission to ECMO insertion, flow rate, turn rate, oxygen flow, peripheral/central, and perioperative treatments, including intra-aortic balloon pump (IABP), percutaneous coronary intervention (PCI), percutaneous transluminal coronary angioplasty (PTCA), and coronary artery bypass graft (CABG). Comorbidities and CCI scores were extracted using the International Classification of Diseases, Ninth Revision, Clinical Modification (ICD-9-CM) before 2016 and the Tenth Revision (ICD-10-CM) in and after 2016. Laboratory data were captured at the time closest to the surgery, always within 24h.

### Statistics

The baseline characteristics and ECMO-related features of the patients who died during admission versus those who survived to discharge were compared using an independent sample t-test for continuous variables or the chi-square test for categorical variables. The trend of the proportions of in-hospital deaths across the study years (2003-2018) was assessed using the Cochran–Armitage test. The trend of LVEF values across ECMO insertion, before discharge and the 180^th^ day after discharge was tested using a generalized estimating equation (with identity linking function and normal distribution), in which the analyzed cohort was restricted to the 403 patients who survived to discharge. LVEF values with the four definitions were categorized into two or three groups, as appropriate. The risk of all-cause mortality between the groups (e.g., LVEF before discharge: <30%, 30-49% and ≥50%) was compared using a log-rank test. Owing to the limited sample size, no covariate adjustment was made. Two-sided *P* values < 0.05 were considered statistically significant. SAS version 9.4 (SAS Institute) was used for statistical analyses.

## Results

### Epidemiological information

The distribution of indications for patients who were weaned and survived to discharge, weaned and died during admission, and failed weaning are listed in **Figure 1**. Both the number of extracorporeal membrane oxygenation (ECMO) procedures and the proportion of in-hospital deaths increased with year (**Figure 2A**). The proportion of postcardiotomy shock decreased over time, while that of AMI and ECPR increased over the years (**Figure 2B**).

**Figure 2.**
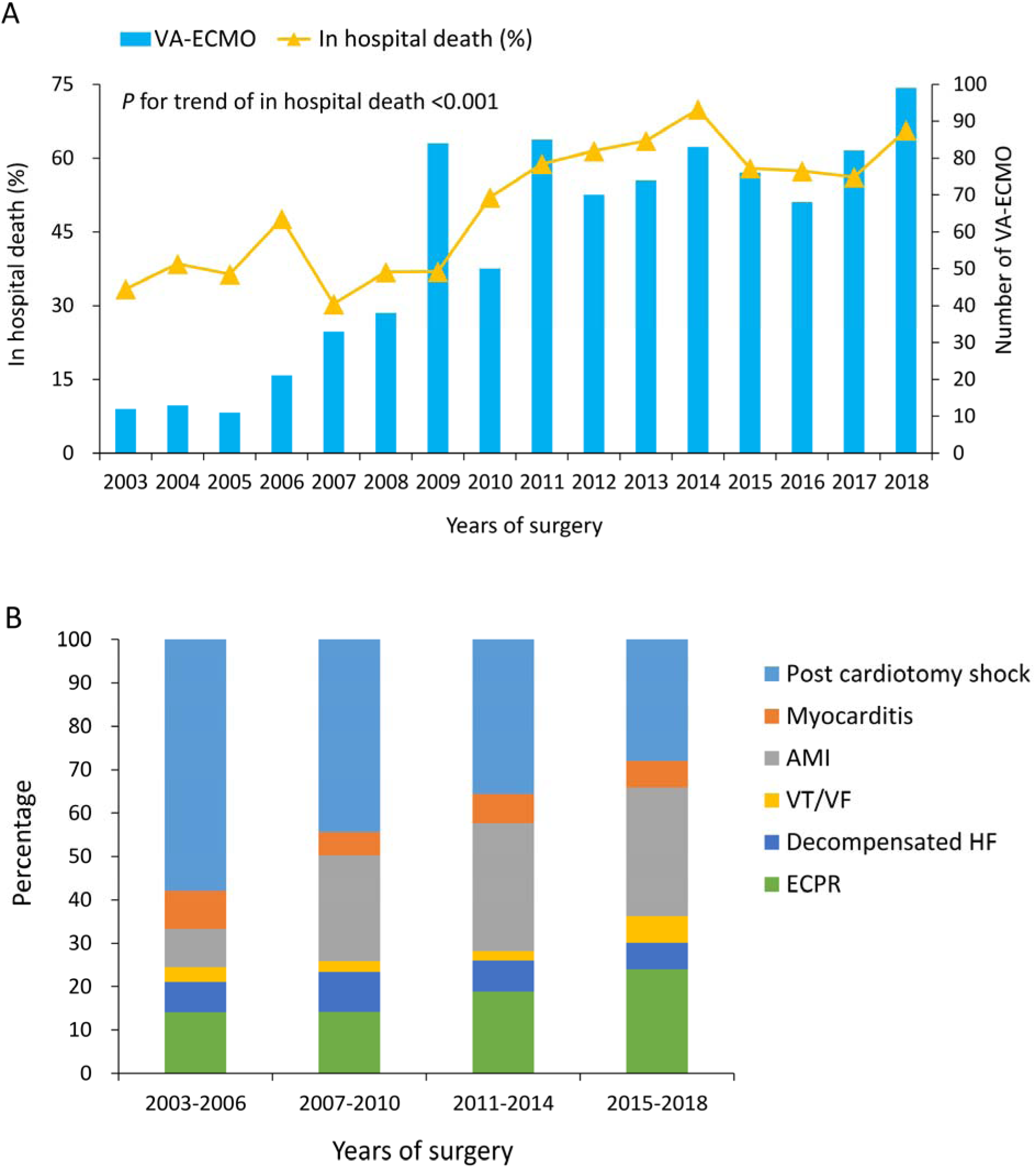
The number of VA-ECMO and proportion of in-hospital death across years (A) and the distribution of indications across the study period (B). VA, venoarterial; ECMO, extracorporeal membrane oxygenation; AMI, acute myocardial infarction; VT, ventricular tachycardia; VF, ventricular fibrillation; HF, heart failure; ECPR, extracorporeal cardiopulmonary resuscitation.

### Baseline characteristics

Of the 899 patients supported with VA-ECMO, the mean age was 58.7 years and 70 percent (n=627) were male. Forty-five percent (n=403) of patients survived to discharge. Compared to patients who died during admission, those who survived to discharge were younger and had a lower prevalence of diabetes, coronary artery disease, peripheral artery disease, myocardial infarction, chronic kidney disease, bleeding events, and malignancy. The CCI score was also significantly lower in patients who survived to discharge. Patients who survived to discharge had lower levels of PaCO^2^, creatinine, liver function markers, and lactic acid but higher levels of PH and platelet count (**Table 1**).

**Table 1.** Baseline characteristics of patients underwent VA-ECMO by the survival status at discharge

| Variable | Available number | Total (n = 899) | In-hospital death (n = 496) | Survivor to discharge (n = 403) | P |
| --- | --- | --- | --- | --- | --- |
| Demographics |  |  |  |  |  |
| Age, year | 899 | 58.7 ± 14.9 | 60.7 ± 14.8 | 56.2 ± 14.7 | <0.001 |
| Male | 899 | 627 (69.7) | 337 (67.9) | 290 (72.0) | 0.192 |
| Body mass index, kg/m<sup>2</sup> | 794 | 25.4 ± 4.6 | 25.5 ± 5.0 | 25.3 ± 4.1 | 0.528 |
| Smoking | 899 | 324 (36.0) | 178 (35.9) | 146 (36.2) | 0.916 |
| Comorbid Conditions |  |  |  |  |  |
| Diabetes | 899 | 295 (32.8) | 182 (36.7) | 113 (28.0) | 0.006 |
| Hypertension | 899 | 472 (52.5) | 276 (55.6) | 196 (48.6) | 0.036 |
| Chronic obstructive pulmonary disease | 899 | 86 (9.6) | 53 (10.7) | 33 (8.2) | 0.206 |
| Liver cirrhosis | 899 | 29 (3.2) | 18 (3.6) | 11 (2.7) | 0.448 |
| Coronary artery disease | 899 | 528 (58.7) | 319 (64.3) | 209 (51.9) | <0.001 |
| Atrial fibrillation | 899 | 152 (16.9) | 84 (16.9) | 68 (16.9) | 0.980 |
| Peripheral artery disease | 899 | 150 (16.7) | 103 (20.8) | 47 (11.7) | <0.001 |
| Cerebrovascular accident | 899 | 83 (9.2) | 54 (10.9) | 29 (7.2) | 0.057 |
| Myocardial infarction | 899 | 195 (21.7) | 128 (25.8) | 67 (16.6) | 0.001 |
| Hyperlipidemia | 899 | 268 (29.8) | 149 (30.0) | 119 (29.5) | 0.868 |
| Chronic kidney disease | 899 | 360 (40.0) | 252 (50.8) | 108 (26.8) | <0.001 |
| Gastrointestinal bleeding | 899 | 121 (13.5) | 82 (16.5) | 39 (9.7) | 0.003 |
| Major bleeding | 899 | 60 (6.7) | 41 (8.3) | 19 (4.7) | 0.034 |

| Variable | Available<br>number | Total<br>(*n* = 899) | In-hospital death<br>(*n* = 496) | Survivor to discharge<br>(*n* = 403) | *P* |
| --- | --- | --- | --- | --- | --- |
| Gout | 899 | 90 (10.0) | 57 (11.5) | 33 (8.2) | 0.101 |
| Malignancy | 899 | 76 (8.5) | 53 (10.7) | 23 (5.7) | 0.008 |
| Heart failure hospitalization | 899 | 109 (12.1) | 60 (12.1) | 49 (12.2) | 0.977 |
| Charlson's Comorbidity Index score | 899 | 3.2 ± 2.7 | 3.8 ± 2.9 | 2.5 ± 2.3 | <0.001 |
| Pre-operative laboratory data |  |  |  |  |  |
| PaCO<sub>2</sub>, mm-Hg | 348 | 38.6 ± 19.1 | 40.7 ± 22.0 | 36.2 ± 14.7 | 0.026 |
| PH | 348 | 7.3 ± 0.2 | 7.2 ± 0.2 | 7.3 ± 0.2 | <0.001 |
| Hemoglobin, g/dL | 882 | 10.9 ± 2.9 | 10.8 ± 3.1 | 11.0 ± 2.6 | 0.250 |
| White blood cell, 10<sup>3</sup>/μL | 874 | 13.6 ± 6.6 | 13.6 ± 7.0 | 13.7 ± 6.0 | 0.786 |
| Platelets, 10<sup>3</sup>/μL | 877 | 163.5 ± 82.0 | 154.8 ± 81.9 | 173.9 ± 81.0 | 0.001 |
| Creatinine, mg/dL | 881 | 2.2 ± 2.1 | 2.4 ± 2.3 | 1.9 ± 1.9 | <0.001 |
| ALT, U/L | 671 | 69.0 [32.0, 205.0] | 77.0 [35.0, 210.0] | 57.0 [29.0, 169.0] | 0.022 |
| AST, U/L | 742 | 102.5 [52.0, 266.0] | 117.0 [59.0, 325.0] | 89.0 [47.5, 198.5] | 0.001 |
| Total bilirubin, mg/dL | 654 | 1.5 ± 1.6 | 1.5 ± 1.8 | 1.4 ± 1.4 | 0.495 |
| Lactic acid, mg/dL | 548 | 95.8 ± 64.9 | 112.6 ± 64.1 | 75.9 ± 60.1 | <0.001 |
| Troponin I, ng/mL | 675 | 22.5 ± 46.9 | 24.1 ± 51.0 | 20.5 ± 41.3 | 0.324 |

Abbreviations: VA, venoarterial; ECMO, extracorporeal membrane oxygenation; PaCO<sub>2</sub>, partial pressure of carbon dioxide; ALT, alanine aminotransferase; AST, aspartate aminotransferase;
Data were presented as frequency (percentage) or mean ± standard deviation.

### ECMO-related features

The most common indications for ECMO were postcardiotomy shock (36.3%), AMI (27%), ECPR (19.4%), decompensated heart failure (7.2%), myocarditis (6.3%), and VT/VF (3.8%). Compared to patients who died during admission, those who survived to discharge were more likely to have postcardiotomy shock (41.7% vs. 31.9%), less likely to have ECPR (12.9% vs. 24.6%), and had more perioperative treatment with PCI/PTCA (21.8% vs. 14.9%) (**Table 2**).

**Table 2.**
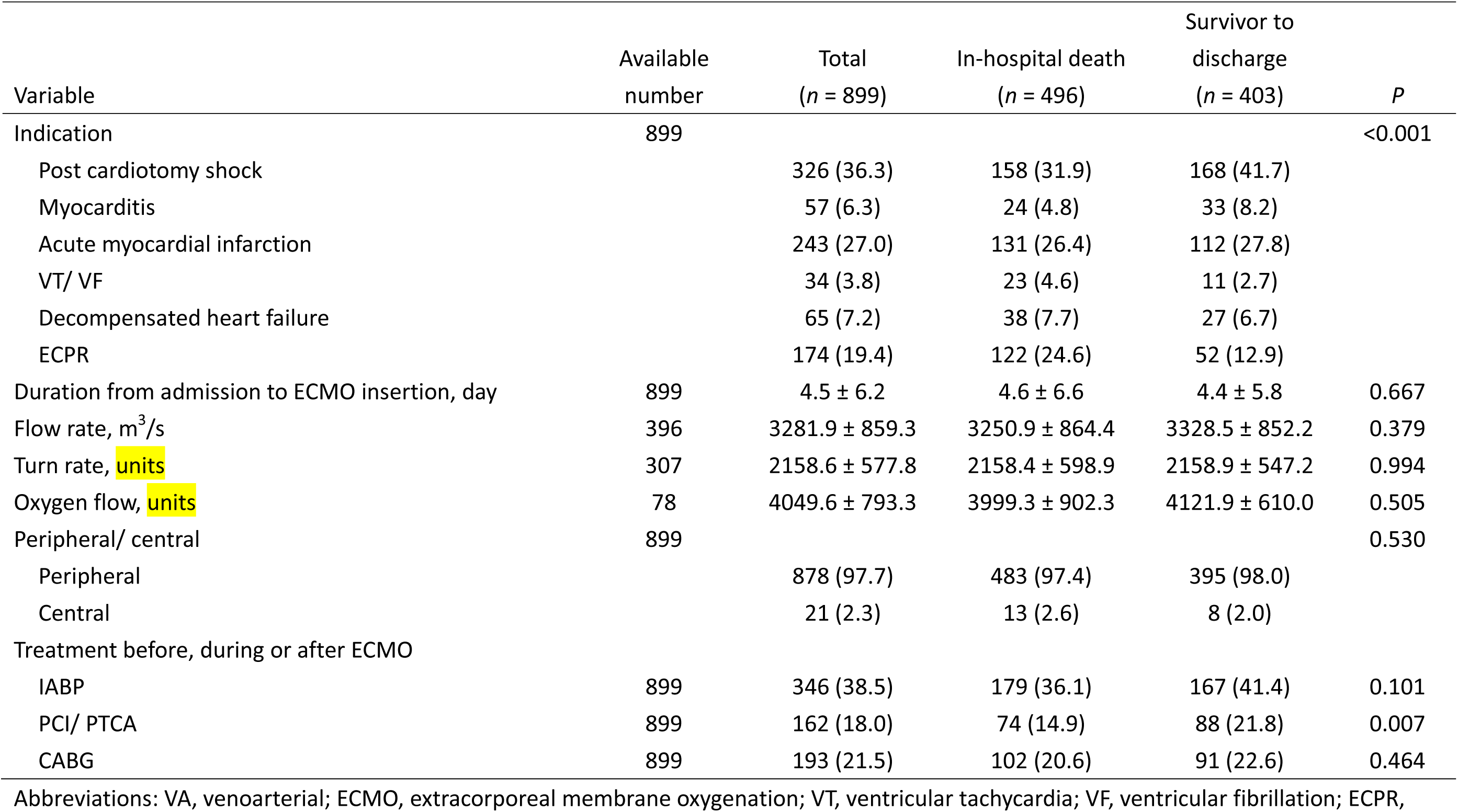
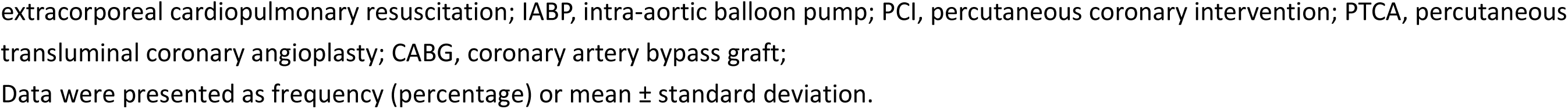
ECMO-related features of patients underwent VA-ECMO by the survival status at discharge

### LVEF and all-cause mortality

The results showed that LVEF significantly improved from ECMO insertion (median: 34%) to before discharge (median: 53%) and the 180^th^ day after discharge (median: 56%) (*P* for trend <0.001; **Figure 3A**). According to the analytical framework (**Figure 3B**), the first analysis showed that the improvement in LVEF from ECMO insertion to before discharge was significantly correlated with a lower mortality rate (*P* for trend <0.001; **Figure 4A**). The second analysis demonstrated that a greater LVEF was significantly correlated with a lower mortality rate (*P* for trend <0.001; **Figure 4B**). The third analysis revealed that the mortality risk of patients who improved (≥10%, n=25) from before discharge to the 180^th^ day after discharge versus those who did not improve (<10%) was not significantly different (*P* = 0.601; **Figure 4C**). Finally, the fourth analysis indicated that a greater LVEF on the 180^th^ day after discharge was significantly correlated with a lower mortality rate (*P* for trend = 0.041; **Figure 4D**).

**Figure 3.**
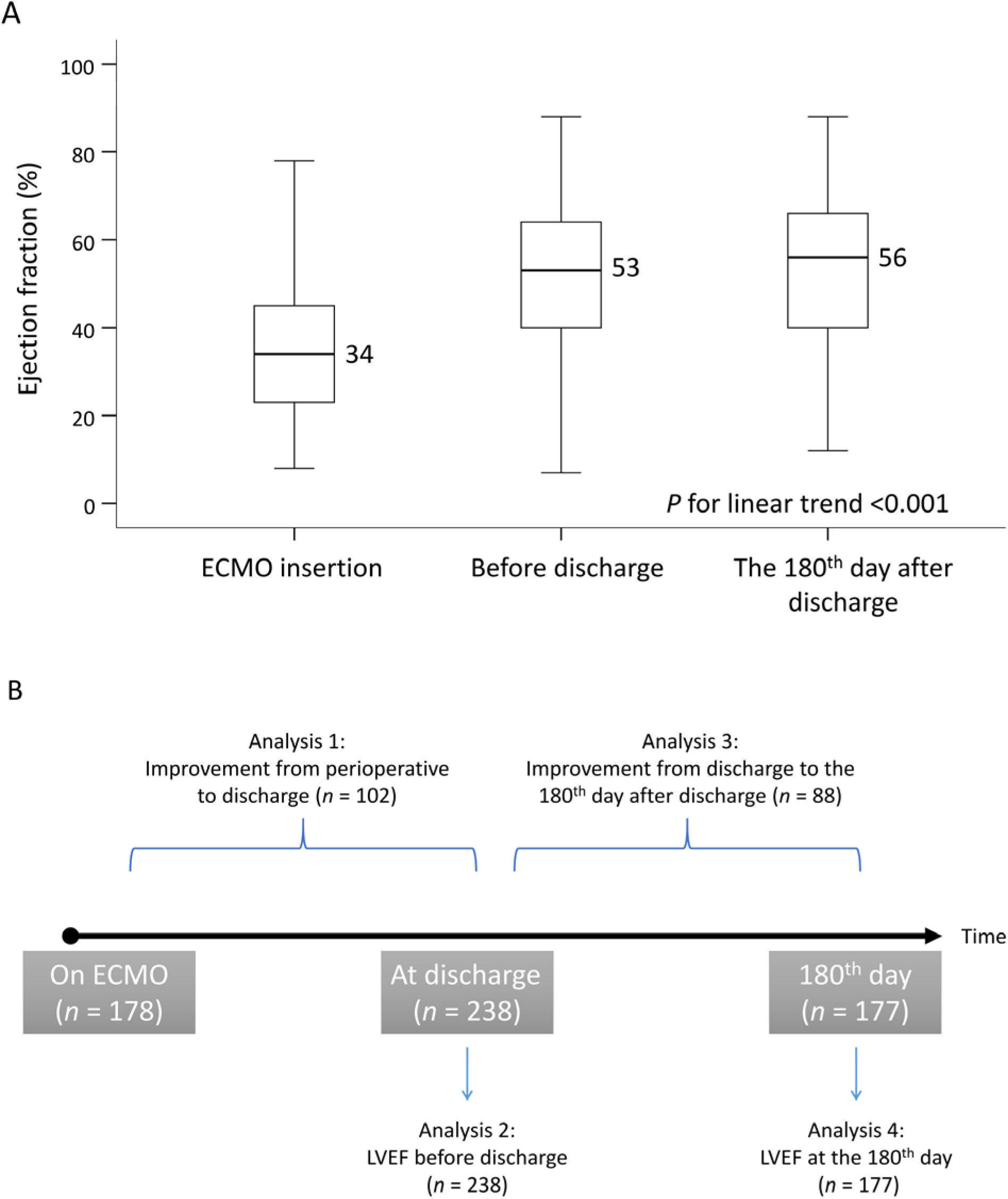
The LVEF values at ECMO insertion, before discharge and the 180^th^ day after discharge (A) and the analytic framework of the study (B). LVEF, left ventricular ejection fraction; ECMO, extracorporeal membrane oxygenation.

**Figure 4.**
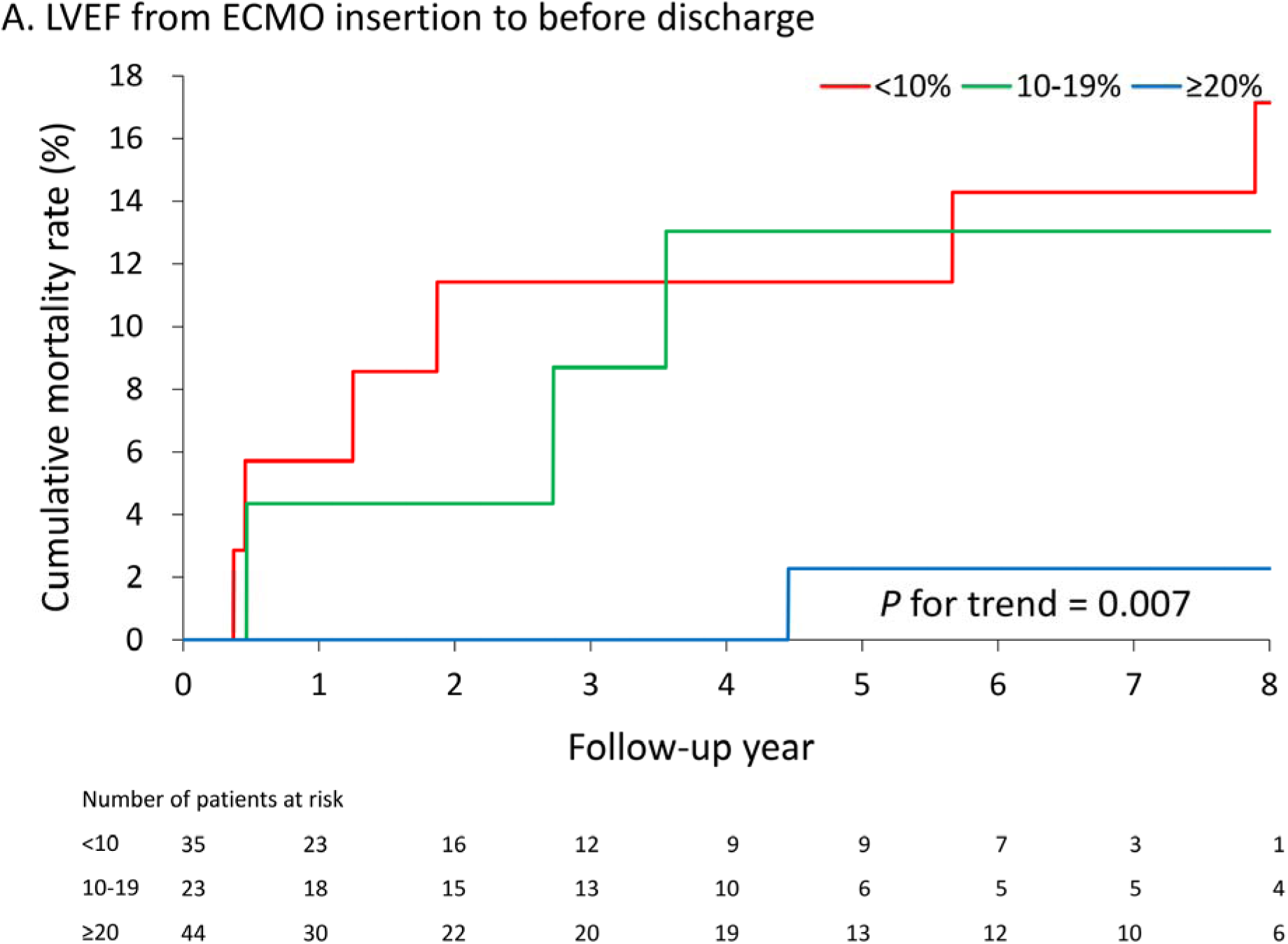
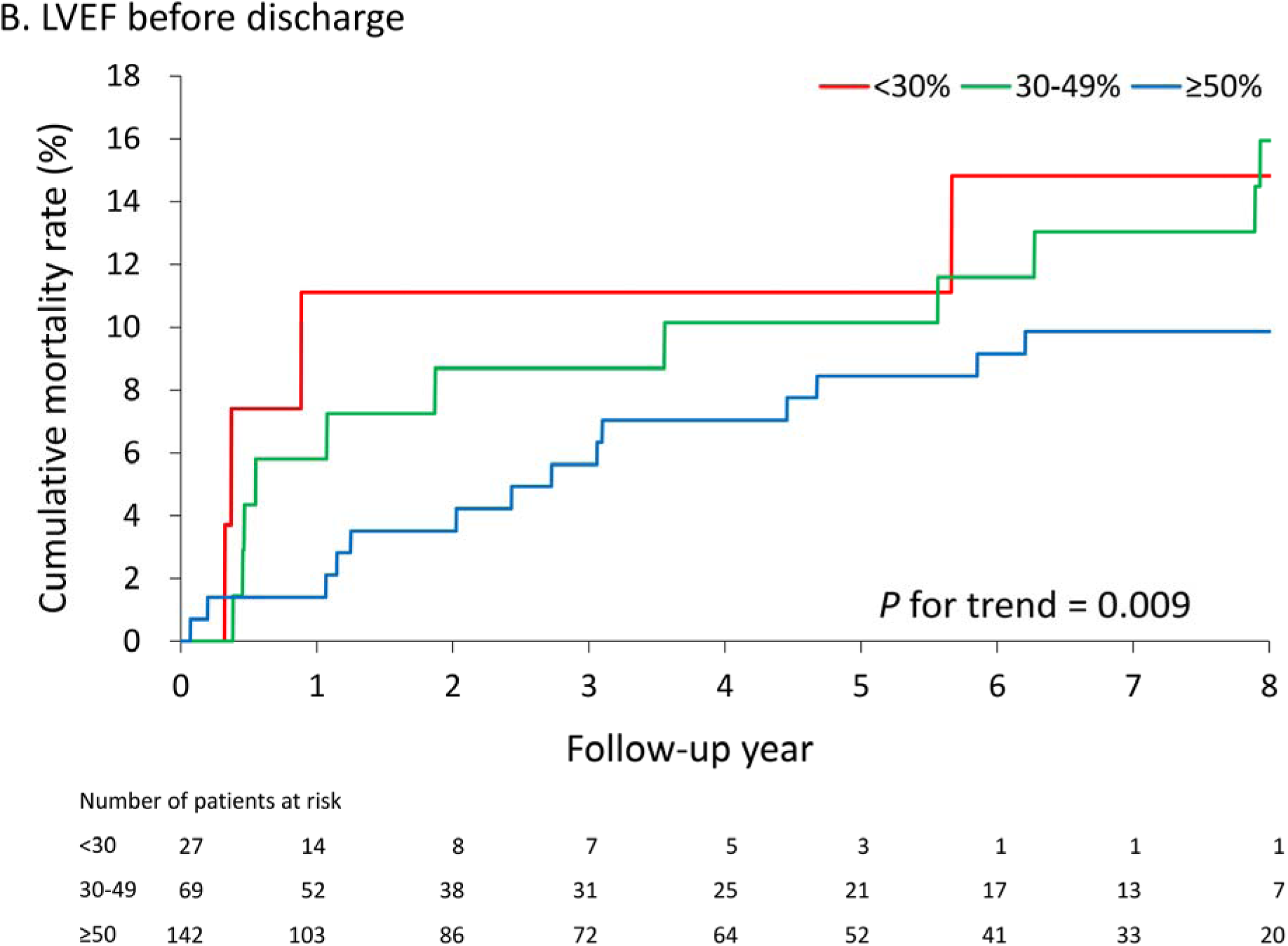
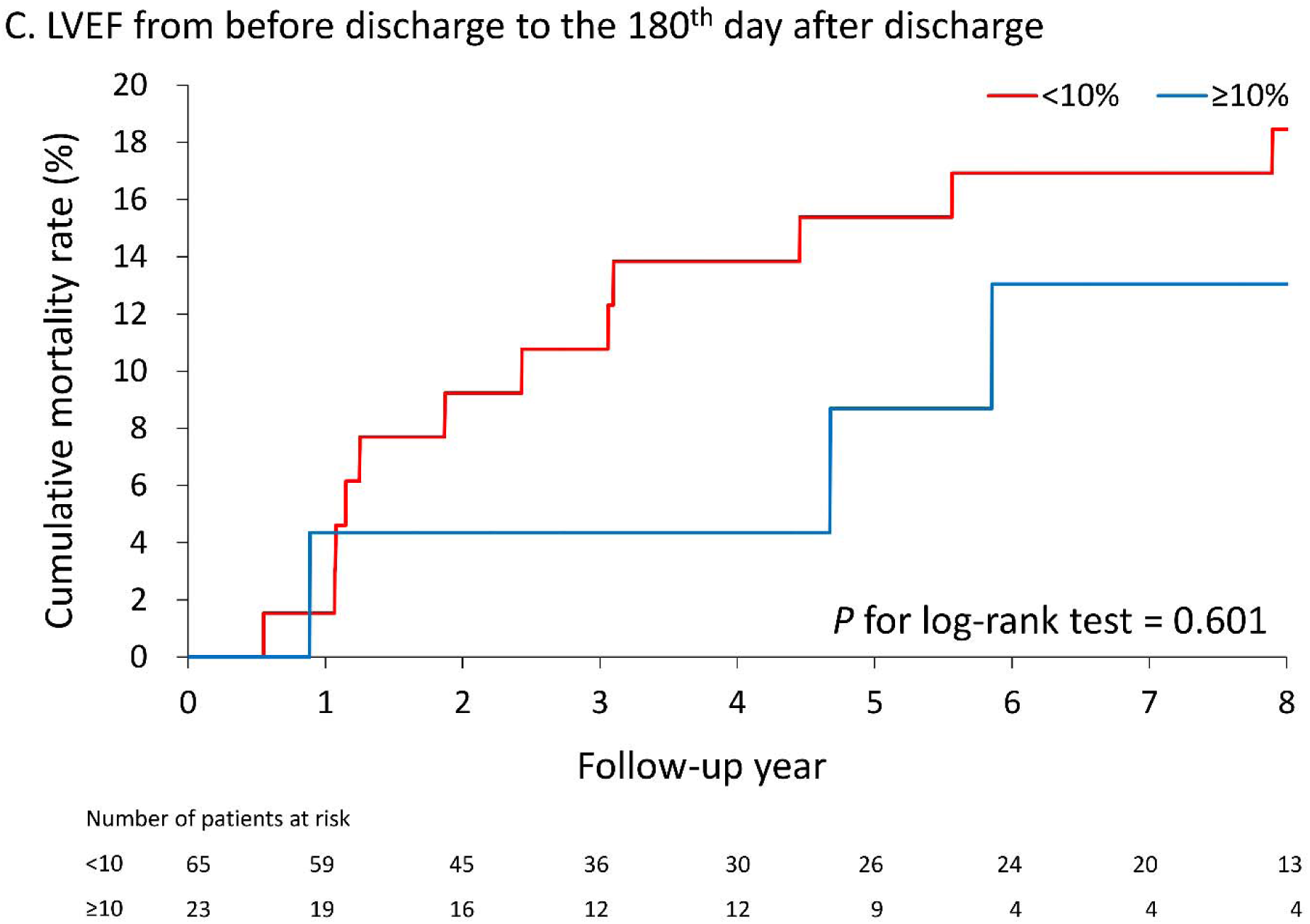
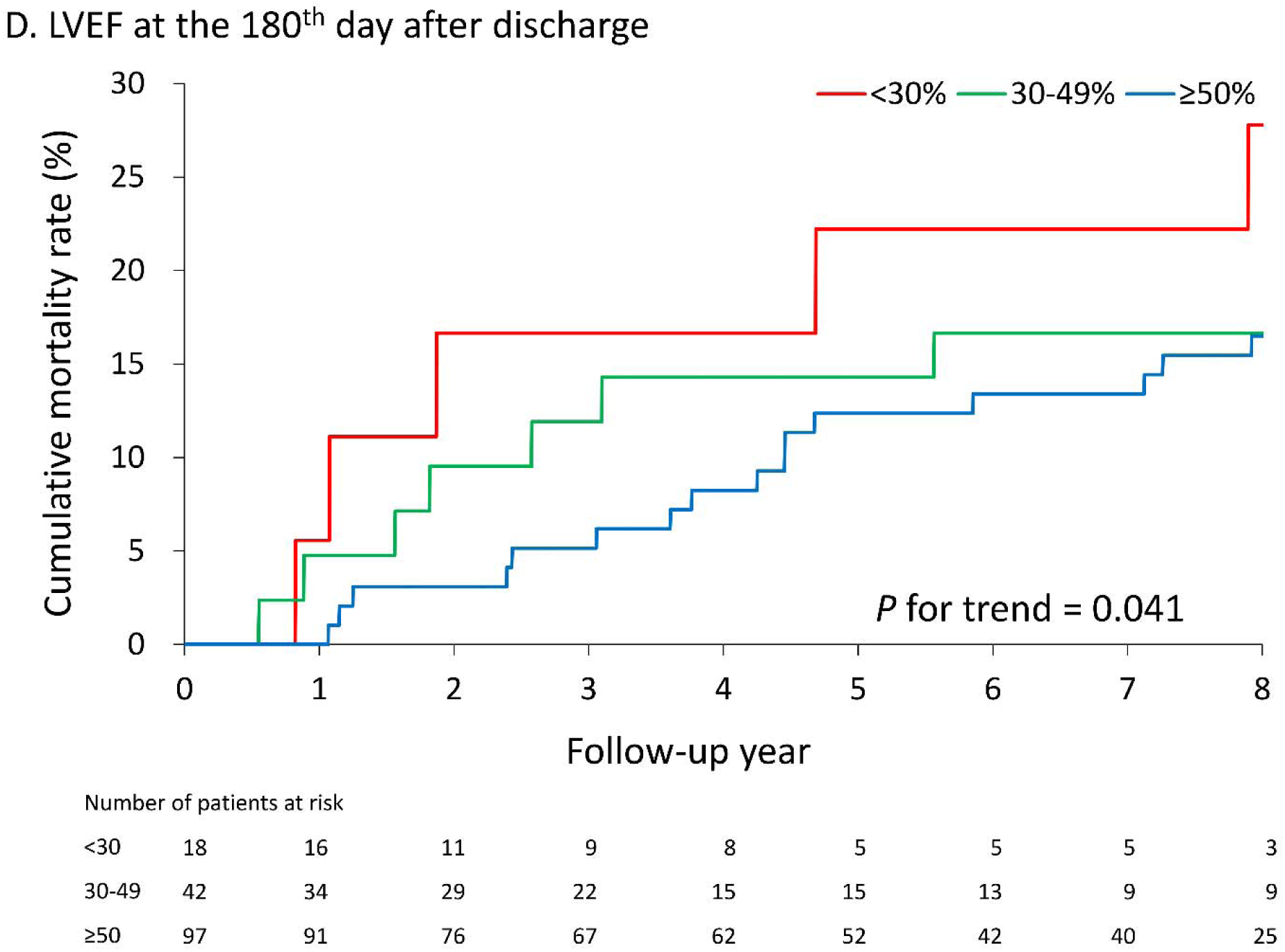
The relationship between risk of all-cause mortality and LVEF with different definitions: the improvement from ECMO insertion to before discharge (A), the LVEF value before discharge (B), before discharge to the 180^th^ day after discharge (C) and LVEF at the 180^th^ day after discharge (D). LVEF, left ventricular ejection fraction; ECMO, extracorporeal membrane oxygenation.

## Discussion

In this multicenter analysis study, we retrospectively evaluated patients with cardiogenic shock (CS) supported with VA-ECMO who successfully survived to hospital discharge. To date, this is the largest multicenter observational retrospective study onf myocardial function in VA-ECMO patients with cardiogenic shock. This is also the first study to provide insights into patients who survived to discharge with their cardiac function data at admission, before discharge, and long-term follow-up with survival analysis. We found that percentile improvement in EF between ECMO placement and successful weaning was significantly associated with lower cumulative mortality. Second, the EF value before discharge was significantly associated with better survival. Lastly, the association of low long-term mortality with EF change from discharge to midterm follow-up and the maximum EF at midterm follow-up was found to be non-significant.

VA-ECMO serves as a temporary bridge (Fukuhara et al. 2018)(Scherer et al. 2009) for myocardial function recovery or further treatment.(Guglin, Zucker et al. 2019)(Ouweneel, D.M. et al. 2016),(Guglin and Burchett 2016) (Squiers et al. 2016). Post-cardiotomy shock develops in around 1% of adult patients who require temporal circulatory mechanical support during or after cardiac surgery with a 63.3% successful weaning rate and 24.8% discharge rate. (Rastan AJ et al. 2010) Chen et al previously showed that despite better outcomes in the first year, compared to non-ECMO groups, the ECMO group showed similar outcome after 1st year follow-up in patients with postcardiotomy shock using the same database in our hospital. In our study, the most common indication for VA-ECMO was postcardiotomy shock, and those who survived until discharge were more likely to have postcardiotomy shock as an indication. (41.7% vs. 31.9%). This finding is in accordance with past research. (Elsharkawy et al 2010) (Chen et al 2017) (Xie et al 2020)

Echocardiography has been shown to be crucial in weaning trials and the assessment of cardiac function recovery. **(Cavarocchi et al 2013)(Aissaoui et al 2015)**. While most studies have focused on identifying mortality predictors at weaning or discharge, there is limited evidence on the short-term and long-term outcomes of VA-ECMO.

In another retrospective study using data from our hospital center, a preoperative LVEF>30% was associated with a lower long-term cumulative mortality rate in patients receiving ECLS for postcardiotomy shock. **(Wu, Meng Yu, et al 2010) Sertic et al** further showed that, severe LV dysfunction (LVEF<30%) was identified to be associated with lower post-weaning survival rate, it was limited to the absolute value of LVEF. Our study is the first to investigate dynamic changes in LV function during ECMO support and from discharge to midterm follow-up. Our study shows that there was an overall improvement in LVEF in patients who survived to midterm follow-up. The major findings of our study suggest that improvements in ejection fraction are strongly associated with favorable long-term survival. An improvement in EF indicates restored myocardial function, and could serve as a potential direct monitoring parameter in complex weaning strategies **(Pappalardo, 2015),** and further treatment since these patients have better long-term survival.

Our study provides further evidence on the impact of EF on long-term survival at different time points. We confirmed the results of past studies that an absolute value of EF<30 before discharge is associated with worse outcomes and found that the EF>50 group fared even better in first-year survival (**Sertic et al 2021; Wu et al 2010**). However, a higher absolute value of the maximum EF at 6 months follow up was not significantly associated with reduced mortality. Improvements in EF from discharge to six months also showed no significance. One possibility to explain this is that improvement in EF from ECMO placement to before discharge or higher EF at weaning plays a more decisive role in deciding mortality, reducing the effect seen on future improvements in EF. Another possibility is that the size of the follow-up group was not sufficiently large to show significance. It is worth noting that percentile improvement of LVEF during VA-ECMO insertion cannot be inferred as the cause of improved long-term mortality, since worse long-term mortality has also been shown to be associated with other predictive factors, such as indications, duration of VA-ECMO support, diabetes, and previous myocardial infarction. (**Sertic, et al. 2021**)**(García-Gigorro et al. 2016**)

Close monitoring and follow-up of discharged patients are therefore warranted to detect groups at risk of first-year failure, or in decisions to arrange ventricular assisting devices or heart transplant in selected candidates. (**Fukuhara et al 2018, Pagani et al 2020**) This result also provides insights for practitioners to view the significant relative improvement of ejection fraction (>20%) as an important clinical hint of promising myocardial function recovery with better long-term survival. At the same time, our results provide physicians with incentives to periodically follow-up myocardial functions, look for alternative therapies to improve EF after discharge, or a future protocol to guide practices based on ultrasound parameters.

## Limitations

This study has several limitations. First, limitations common to its retrospective and observational nature may be relevant, including the possibility of a selection bias. Further analyses are required for patients who are bridged to VADs and heart transplants and for those with indications not of interest. Confounding factors that could affect survival other than directly changing myocardial function render the relationship between survival and ejection fraction relative rather than causative. Second, the patient group size in our series could have affected the significance of our results, especially in the analyses involving missing midterm and long-term EF data due to loss of follow-up or changes in electronic records. Finally, there is a limitation in extending the results to other countries owing to the higher ECPR usage in Taiwan (**Hsu et al. 2012**), which lacks records of baseline EF at presentation to the emergency room and lower numbers of VADs and heart transplants.

## Conclusion

Ejection fraction monitoring at ECMO insertion and before discharge can inform physicians regarding patients’ long-term outcomes. EF percentile improvement from insertion to hospital discharge and a higher absolute EF value before discharge were associated with lower long-term mortality rates. Studies of other cardiac echocardiography parameters using long-term follow-up data should be performed in future research to validate these findings.

## Funding statement

This work was supported by a grant from Chang Gung Memorial Hospital, [Taiwan CORPG3M0371, CFRPG3M0011, CMRPG3L0101, CMRPG3L0102, CMRPG3L0103 and BMRPD95(SWC)]. This work was also supported by the Ministry of Science and Technology grant [Most 110-2314-B-182A-114 (SWC].

## Disclosures

No conflicts of interest for all authors.

## Data Availability

all the data can be requested be email

## Acknowledgments

This study was based on data from the NHIRD, which was provided by the NHI Administration, Ministry of Health and Welfare of Taiwan, and managed by the National Health Research Institutes of Taiwan. However, the interpretation and conclusions contained in this paper represent only the opinions of the authors. The authors would like to thank and acknowledge the support of the Maintenance Project of the Center for Big Data Analytics and Statistics (Grant CLRPG3D0049) at Chang Gung Memorial Hospital for help with statistical consultation and data analysis. The authors also thank Alfred Hsing-Fen Lin and Zoe Ya-Jhu Syu, Raising Statistics Consultant Inc., for their assistance with the statistical analysis. This manuscript was edited by Wallace Academic Editing.

## Notes

### Competing Interest Statement

The authors have declared no competing interest.

### Author Declarations

Informed consent was obtained from the institutional review board of CGMH (approval number: 202100124B)

